## Supplementary Materials for "Genomic risk prediction for breast cancer in older women"

**Figure S1: Principal component (PC) analysis of the ASPREE cohort compared with the 1,000 Genome Project.**

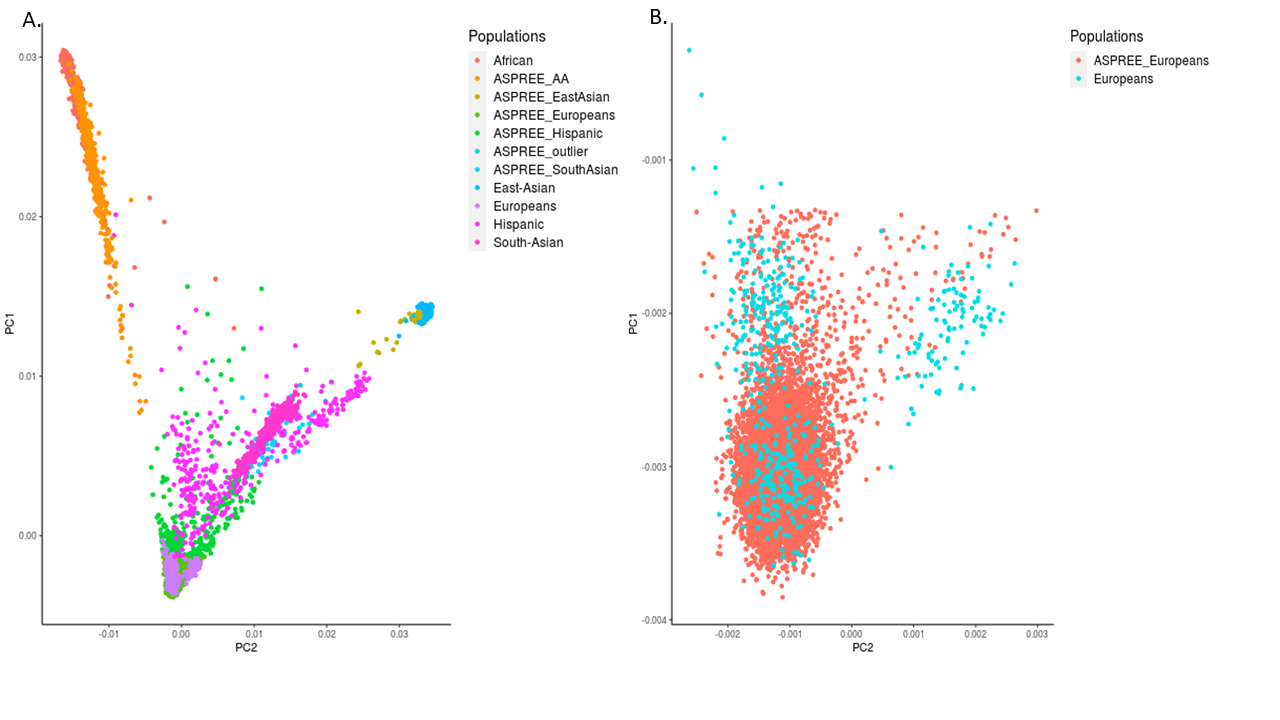

**A.** PC plot of all genoptyped ASPREE participants mapped against the 1,000 Genome population groups (Europeans, South Asians, East Asians, African American and Hispanics). (ASPREE_AA = ASPREE participants of African American descent).

**B.** PC plot of European ASPREE genotyped participants included in the PRS study mapped against the 1,000 Genome European population.

### **Figure S2**

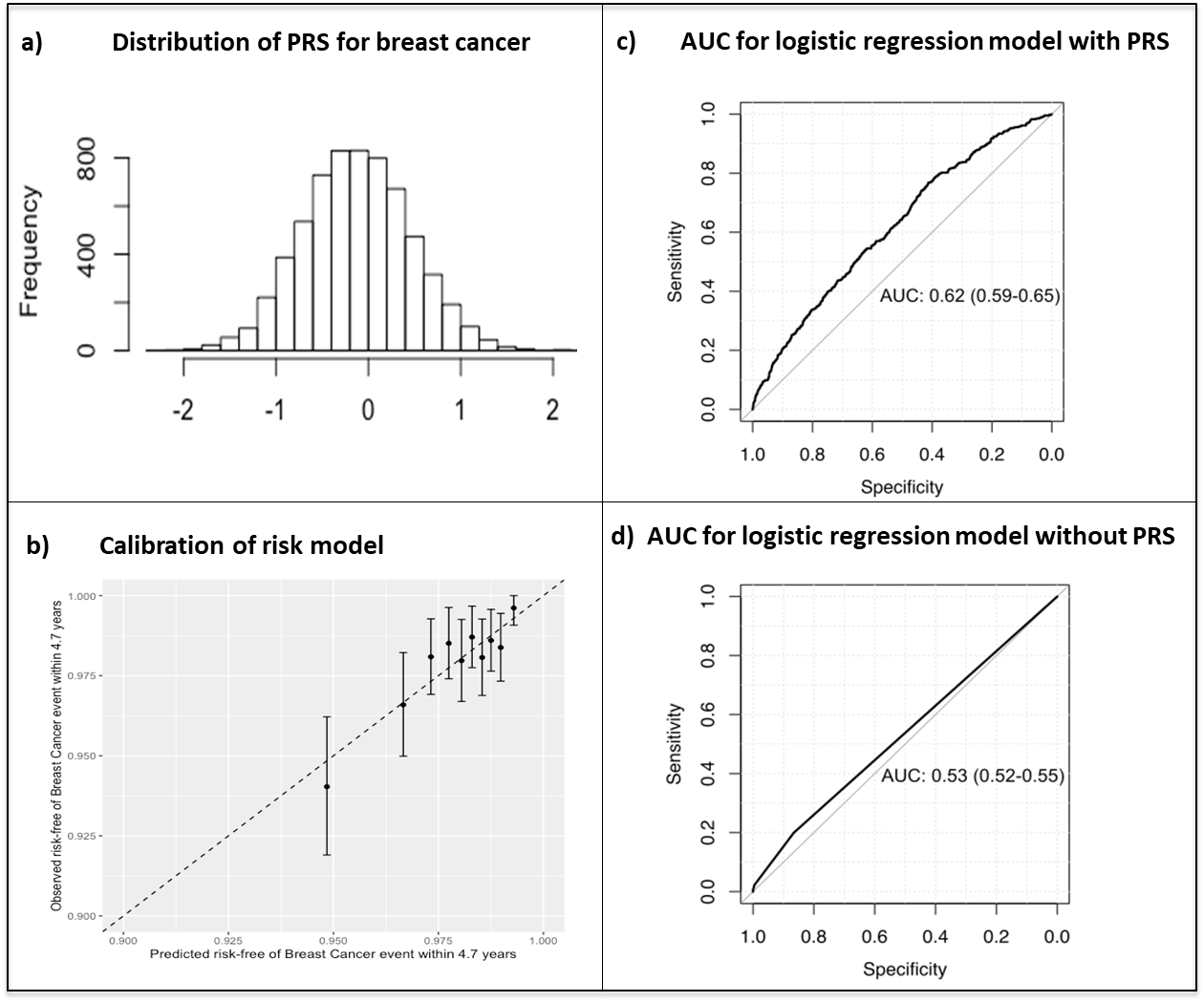

**Table S1: Table of pathogenic variants (PVs) detected in breast cancer susceptibility genes (*BRCA1, BRCA2, PALB2, CHECK2, ATM*) in 6,339 older women.** Variants with ‘pathogenic’ or ‘likely pathogenic’ ClinVar annotation and/or high-confidence predicted loss-of-function in coding regions were curated following ACMG/AMP Standards and Guidelines for the Interpretation of Sequence Variants, including review by two or more laboratory scientists and a clinical geneticist. Analysis was restricted to single nucleotide variants and small insertions/deletions.

| **Location (GRCh37)** | **Gene** | **REF** | **ALT** | **Consequence** | **HGVS** | **rsID** | **Curation call** | **ASPREE Allele Count** | **ASPREE MAF (N6339)** | **Gnomad NFE MAF** |
| --- | --- | --- | --- | --- | --- | --- | --- | --- | --- | --- |
| chr17:41276044 | *BRCA1* | ACT | A | Deletion | NM_007294.3(BRCA1):c.68_69delAG, p.(Glu23ValfsTer17) | rs80357914 | Pathogenic | 1 | 0.000158 | 0.000097 |
| chr17:41245528 | *BRCA1* | GT | G | Deletion | NM_007294.3(BRCA1):c.2019delA, p.(Glu673AspfsTer28) | rs80357626 | Pathogenic | 1 | 0.000158 | 0 |
| chr17:41245528 | *BRCA1* | GAA | G | Deletion | NM_007294.3(BRCA1):c.798_799delTT, p.(Ser267LysfsTer19) | rs80357724 | Pathogenic | 1 | 0.000158 | 0.000085 |
| chr13:32893291 | *BRCA2* | G | T | stop_gained | NM_000059.3(BRCA2):c.145G>T, p.(Glu49Ter) | rs80358435 | Pathogenic | 1 | 0.000158 | 0 |
| chr13:32912964 | *BRCA2* | TGAAA | T | Deletion | NM_000059.3(BRCA2):c.4478_4481delAAAG, p.(Glu1493ValfsTer10) | rs80359454 | Pathogenic | 2 | 0.00032 | 0.000035 |
| chr13:32929057 | *BRCA2* | TTC | T | Deletion | NM_000059.3(BRCA2):c.7069_7070delCT, p.(Leu2357ValfsTer2) | rs80359636 | Pathogenic | 2 | 0.000158 | 0.000053 |
| chr13:32912337 | *BRCA2* | CTG | C | Deletion | NM_000059.3(BRCA2):c.3847_3848delGT, p.(Val1283LysfsTer2) | rs80359405 | Pathogenic | 1 | 0.000158 | 0.000114 |
| chr13:32954180 | *BRCA2* | CG | C | Deletion | NM_000059.3(BRCA2):c.9157delG, p.(Glu3053SerfsTer9) | rs80359750 | Pathogenic | 1 | 0.000158 | 0 |
| chr13:32893460 | *BRCA2* | T | G | stop_gained | NM_000059.3(BRCA2):c.314T>G, p.(Leu105Ter) | rs80358561 | Pathogenic | 1 | 0.000158 | 0 |
| chr13:32914401 | *BRCA2* | C | A | stop_gained | NM_000059.3(BRCA2):c.5909C>A, p.(Ser1970Ter) | rs80358824 | Pathogenic | 1 | 0.000158 | 0 |
| chr13:32912171 | *BRCA2* | CTG | C | Deletion | NM_000059.3(BRCA2):c.3680_3681delTG, p.(Leu1227GlnfsTer5) | rs80359395 | Pathogenic | 1 | 0.000158 | 0 |
| chr11:108199929 | *ATM* | T | G | SNV | NM_000051.3(ATM):c.7271T>G (p.Val2424Gly) | rs28904921 | Pathogenic | 1 | 0.000158 | 0.000079 |
| chr11:108236203 | *ATM* | C | T | SNV | NM_000051.3(ATM):c.9139C>T (p.Arg3047Ter) | rs121434219 | Pathogenic | 1 | 0.000158 | 0 |
| chr11:108143258 | *ATM* | G | A | SNV | NM_000051.3(ATM):c.3078-1G>A | rs750663117 | Pathogenic | 1 | 0.000158 | 0.000018 |
| chr11:108224608 | *ATM* | G | A | splice_donor | NM_000051.3(ATM):c.8786+1G>A | rs17174393 | Pathogenic | 1 | 0.000158 | 0.000035 |
| chr11:108213987 | *ATM* | G | A | stop_gained | NM_000051.3(ATM):c.8307G>A (p.Trp2769Ter) | rs778269655 | Pathogenic | 1 | 0.000158 | 0.000009 |
| chr11:108203621 | *ATM* | C | T | stop_gained | NM_000051.3(ATM):c.7921C>T (p.Gln2641Ter) | rs769523686 | Likely pathogenic | 1 | 0.000158 | 0.000009 |
| chr11:108124740 | *ATM* | C | T | stop_gained | NM_000051.3(ATM):c.2098C>T (p.Gln700Ter) | rs786202743 | Pathogenic | 1 | 0.000158 | 0 |
| chr11:108213981 | *ATM* | TGAATGG  TGCACAG | T | Deletion | NM_001330368.2(C11orf65):c.641-34196_641-34184del | rs786202318 | Pathogenic | 1 | 0.000158 | 0 |
| chr11:108225581 | *ATM* | ACT | A | Deletion | NM_000051.4(ATM):c.8831_8832CT[1] (p.Leu2945fs) | rs786203030 | Pathogenic | 1 | 0.000158 | 0.000009 |
| chr11:108155007 | *ATM* | AG | A | Deletion | NM_001351834.2(ATM):c.3802del (p.Glu1267_Val1268insTer) | rs587779834 | Pathogenic | 1 | 0.000158 | 0.000062 |
| chr11:108121752 | *ATM* | CAG | C | Deletion | NM_001351834.2(ATM):c.1562_1563GA[1] (p.Glu522fs) | rs587779817 | Pathogenic | 1 | 0.000158 | 0.000106 |
| chr11:108196890 | *ATM* | CAG | C | Deletion | NM_000051.4(ATM):c.6914_6915AG[1] (p.Leu2307fs) | rs878853535 | Pathogenic | 1 | 0.000158 | 0 |
| chr11:108155055 | *ATM* | TA | T | Deletion | NM_000051.3(ATM):c.3850del (p.Thr1284fs) | rs876660865 | Pathogenic | 1 | 0.000158 | 0 |
| chr11:108121479 | *ATM* | CTG | C | Deletion | NM_001351834.2(ATM):c.1288_1289TG[1] (p.Cys430_Glu431delinsTer) | rs587781598 | Pathogenic | 1 | 0.000158 | 0 |
| chr11:108213970 | *ATM* | AGT | A | Deletion | NM_000051.3(ATM):c.8292_8293del (p.Ser2764fs) | rs879254036 | Pathogenic | 1 | 0.000158 | 0.000009 |
| chr11:108129762 | *ATM* | C | A | stop_gained | NM_000051.3(ATM):c.2426C>A (p.Ser809Ter) | rs730881348 | Pathogenic | 1 | 0.000158 | 0 |
| chr22:29121230 | *CHEK2* | C | T | splice_donor | NM_007194.4(CHEK2):c.444+1G>T | rs121908698 | Pathogenic | 1 | 0.000158 | 0.000167 |
| chr22:29120965 | *CHEK2* | CT | C | Deletion | NM_007194.4(CHEK2):c.591del (p.Val198fs) | rs587782245 | Pathogenic | 1 | 0.000158 | 0.000035 |
| chr22:29091226 | *CHEK2* | TA | T | Deletion | NM_007194.4(CHEK2):c.1263del (p.Ser422fs) | rs587780174 | Pathogenic | 4 | 0.000631 | 0.000088 |
| chr22:29121269 | *CHEK2* | AT | A | Deletion | NM_007194.4(CHEK2):c.405del (p.Lys135fs) | rs730881699 | Pathogenic | 1 | 0.000158 | 0 |
| chr16:23641149 | *PALB2* | A | AT | Duplication | NM_024675.3(PALB2):c.2325dup (p.Phe776fs) | rs876659997 | Pathogenic | 1 | 0.000158 | 0 |
| chr16:23637576 | *PALB2* | TAA | T | Deletion | NM_024675.3(PALB2):c.2727_2728del (p.Thr911fs) | rs730881869 | Pathogenic | 1 | 0.000158 | 0.000009 |
| chr16:23632683 | *PALB2* | C | T | stop_gained | NM_024675.3(PALB2):c.3113G>A (p.Trp1038Ter) | rs180177132 | Pathogenic | 4 | 0.000631 | 0.000114 |

### **Table S2: Categorical net reclassification improvement after adding Polygenic Risk Score to the conventional model to predict 4.7-years risk of breast cancer.**

|  |  | Conventional Model + Polygenic Risk Score | | | |
| --- | --- | --- | --- | --- | --- |
|  | Conventional Model | < 1% | 1 to 2.99% | ≥ 3% | Total No.  of participants |
| Breast Cancer Event | < 1% | 1 | 1 | 0 | 2 |
|  | 1 to 2.99% | 3 | 56 | 17 | 76 |
|  | ≥ 3% | 0 | 2 | 30 | 32 |
|  | Total No.  of participants | 4 | 59 | 47 | 101 |
| No Breast Cancer Event | < 1% | 162 | 58 | 0 | 220 |
|  | 1 to 2.99% | 333 | 1726 | 185 | 2244 |
|  | ≥ 3% | 0 | 103 | 238 | 341 |
|  | Total No.  of participants | 495 | 1887 | 423 | 2805 |

**Table S3: Receptor Subtypes**

| **Receptor Subtype** | **Incident Cases** |
| --- | --- |
| ER+ | 9 |
| PR+ | 2 |
| HER2+ | 4 |
| ER+/PR+ | 74 |
| ER+/HER2+ | 3 |
| ER+/PR+/HER2+ | 5 |
| Triple Negative | 6 |

**Table S4: Association of rare pathogenic variants (PVs) and a polygenic risk score (PRS) with prevalent breast cancer risk in 6,339 older women.** We used a logistic regression model including family history of BC to report the Odds Ratio (OR) of rare PVs and the PRS for prevalent BC risk, based on BC cases diagnosed before the time of enrolment (475 self-reported cases).

|  | **PRS as Continuous Variable** | | | **PRS as Categorical Variable** | | |
| --- | --- | --- | --- | --- | --- | --- |
|  | **Odds**  **Ratio** | **95% CI** | **p-value** | **Odds**  **Ratio** | **95% CI** | **p-value** |
| **Family History of**  **Breast Cancer*** | 1.41 | (1.10; 1.80) | 0.006 | 1.43 | (1.12; 1.83) | 0.004 |
| **Pathogenic Variants**  (N41 carriers) | 4.69 | (2.21; 9.27) | <0.001 | 4.64 | (2.19; 9.15) | <0.001 |
| **Polygenic Score**  (per standard deviation) | 1.47 | (1.34; 1.61) | <0.001 |  |  |  |
| **Low PRS**  0-20% (Q1) |  | | | Reference | | |
| **Moderate PRS**  21-80% (Q2,3,4) |  |  |  | 2.12 | (1.56; 2.94) | <0.001 |
| **High PRS**  81-100% (Q5) |  |  |  | 3.16 | (2.26; 4.49) | <0.001 |

*Family history in first-degree blood relative (mother, sibling or child)

PRS = Polygenic risk score, CI = Confidence interval

**Table S5: Per-gene odds ratios (ORs) for prevalent BC risk in pathogenic variant carriers**

| **Gene** | **Number of female carriers** | **OR for BC** | **95% CI** | **p.value** |
| --- | --- | --- | --- | --- |
| *BRCA1* | 3 | 18.5 | 1.72 - 404.61 | 1.85E-02 |
| *BRCA2* | 10 | 4.03 | 0.81 - 15.58 | 5.59E-02 |
| *ATM* | 16 | 1.95 | 0.30 - 7.14 | 3.81E-01 |
| *PALB2* | 6 | 7.18 | 0.97 - 37.93 | 2.54E-02 |
| *CHEK2* | 7 | 4.42 | 0.58 - 22.12 | 9.22E-02 |
| **Gene groups** |  |  |  |  |
| *BRCA1/2* | 13 | 6.09 | 1.77 - 19.08 | 2.29E-03 |
| *non-BRCA1/2* | 29 | 3.33 | 1.19 - 7.92 | 1.14E-02 |

**Table S6: Association of rare pathogenic variants (PVs) and a polygenic risk score (PRS) with prevalent breast cancer risk, stratified by age at diagnosis.**

|  | **Diagnosis Age < 50yrs**  n=60 cases | | | **Diagnosis Age 50+ years**  n=415 cases | | |
| --- | --- | --- | --- | --- | --- | --- |
|  | **Odds**  **Ratio** | **95% CI** | **p-value** | **Odds**  **Ratio** | **95% CI** | **p-value** |
| **Family History of**  **Breast Cancer *** | 2.09 | (1.12; 3.69) | 0.004 | 1.34 | (1. 20; 1.74) | 0.03 |
| **Pathogenic Variants**  (N41 carriers) | 9.79 | (2.29; 28.87) | 0.02 | 3.91 | (1.64; 8.31) | <0.0001 |
| **No. of Children ǂ** | Not available | | | 0.79 | (0.71; 0.88) | <0.0001 |
| **Polygenic Score**  (per std dev) | 1.39 | (1.08; 1.80) | 0.01 | 1.47 | (1.33; 1.63) | <0.0001 |

HR = Hazard ratio, CI = Confidence interval, std dev = Standard deviation

*Family history in first-degree blood relative (mother, sibling or child)

**ǂ** Assumes all participants had children by age 50.

**Table S7: Chi-squared test (χ2=1.97, df=2, P=0.37)**

| **PRS Group** | **Low (1-20)** | **Med (21-80)** | **High (81-100)** |
| --- | --- | --- | --- |
| Monogenic case | 2 | 8 | 1 |
| Monogenic control | 7 | 15 | 8 |
